# Choosing the right tool for the job: A comprehensive assessment of serological assays for SARS-CoV-2 as surrogates for authentic virus neutralization

**DOI:** 10.1101/2021.04.14.21255399

**Authors:** Nicholas Wohlgemuth, Kendall Whitt, Sean Cherry, Ericka Kirkpatrick Roubidoux, Chun-Yang Lin, Kim J. Allison, Ashleigh Gowen, Pamela Freiden, E. Kaitlynn Allen, St. Jude Investigative Team, Aditya H. Gaur, Jeremie H. Estepp, Li Tang, Tomi Mori, Diego R. Hijano, Hana Hakim, Maureen A. McGargill, Florian Krammer, Michael A. Whitt, Joshua Wolf, Paul G. Thomas, Stacey Schultz-Cherry

**Author notes:** Corresponding author: Stacey-Schulz-Cherry.

## Abstract

Severe acute respiratory syndrome coronavirus 2 (SARS-CoV-2) emerged in late 2019 and has since caused a global pandemic resulting in millions of cases and deaths^1-4^. Diagnostic tools and serological assays are critical for controlling the outbreak^5-7^, especially assays designed to quantitate neutralizing antibody levels, considered the best correlate of protection^8-11^. As vaccines become increasingly available^12^, it is important to identify reliable methods for measuring neutralizing antibody responses that correlate with authentic virus neutralization but can be performed outside of biosafety level 3 (BSL3) laboratories. While many neutralizing assays using pseudotyped virus have been developed, there have been few studies comparing the different assays to each other as surrogates for authentic virus neutralization^9,10,13,14^. Here we characterized three enzyme-linked immunosorbent assays (ELISAs) and three pseudotyped VSV virus neutralization assays and assessed their concordance with authentic virus neutralization. The most accurate assays for predicting authentic virus neutralization were luciferase and secreted embryonic alkaline phosphatase (SEAP) expressing pseudotyped virus neutralizations, followed by GFP expressing pseudotyped virus neutralization, and then the ELISAs.

---

Numerous serological assays have been developed to quantitate severe acute respiratory syndrome coronavirus 2 (SARS-CoV-2) antibody responses to determine total antibody concentration and neutralization activity. Neutralization assays can be performed with authentic virus or a pseudotyped virus expressing the SARS-CoV-2 spike (S) protein on its surface and a marker to measure infection of cells. The clear advantage of a pseudotyped virus is safety, as these studies can be performed in standard BSL2 laboratories. Another advantage is that results using pseudotyped virus can be obtained sooner, typically less than 24 hours whereas with authentic virus, plaque reduction-based neutralization assays take 2-3 days. A third advantage of using pseudotypes is flexibility. Pseudotypes expressing spike variants can be generated easily once the sequence is known since all that is needed is a plasmid that expresses the variant of interest. One disadvantage of the pseudotyped virus neutralization assay is the pseudotyped viruses lack all but the spike protein from SARS-CoV-2, meaning they can only be neutralized by spike specific antibodies, and the clustering of proteins may not be representative of authentic virus particles. Yet, few studies have demonstrated whether the neutralization dose 50% (ND_50_), the dilution at which 50% of virus will be neutralized, differs between pseudotyped virus detection platforms and, importantly, how they compare to authentic virus. To fill this gap in knowledge, we compared SARS-CoV-2 antibody titers in plasma from 24 PCR-positive individuals and 10 PCR negative individuals by enzyme-linked immunosorbent assay (ELISA), pseudotyped virus neutralization assay, and authentic virus neutralization. Adult participants were enrolled in the prospective, adaptive cohort study of St. Jude Children’s Research Hospital employees “St. Jude Tracking of Viral and Host Factors Associated with COVID-19” (SJTRC, clinicaltrials.gov #NCT04362995) beginning in April of 2020. SJTRC was approved by the St. Jude Internal Review Board, and all participants provided written informed consent in a manner consistent with institutional policies. Cohort characteristics are provided in Supplemental Table 1. Samples were collected from PCR-positive individuals a median 33 days following diagnosis (interquartile range: 25.75-48.25) between April and August 2020.

The ELISAs included in the comparison detect antibodies to SARS-CoV-2 spike protein, nucleocapsid protein (N), or the receptor-binding domain (RBD) of the spike protein as described^15^. Briefly, plasma samples were diluted 1:50 for RBD and N ELISAs and results expressed as the ratio of the optical density (OD) from the sample over the negative control (a known negative, pre-pandemic plasma sample), which is common practice. To determine spike titers, plasma was diluted 1:100 to 1:8100 and an area under the curve (AUC) analysis performed. The pseudotyped virus platform was a vesicular stomatitis virus (VSV) glycoprotein (G) knockout VSV expressing full-length SARS-CoV-2 spike protein (VSV-ΔG-S) from the Wuhan-Hu-1 strain with three different reporter genes: green fluorescence protein (GFP), luciferase (Luci.) and secreted alkaline phosphatase (SEAP). Authentic virus neutralization studies were performed under BSL3+ conditions with the 2019n-CoV/USA_WA1/2020 strain obtained from BEI Resources.

All PCR positive participants had ELISA titers to RBD, N, and spike, although the titers differed (Table 1). The average RBD ratio for the positive participants was 16.45 (95% CI: 14.53 –18.36) and 1.62 (95% CI: 1.33 – 1.91) for negative participants while the average N ratios were 9.20 (95% CI: 7.69 – 11.11) for positive participants and 1.40 (95% CI: 0.79 – 2.01) for negative participants. Spike AUC average was 6.44 (95% CI: 5.04 – 7.85) for the positive samples (the spike ELISA was not performed on negative samples).

**Table 1:**
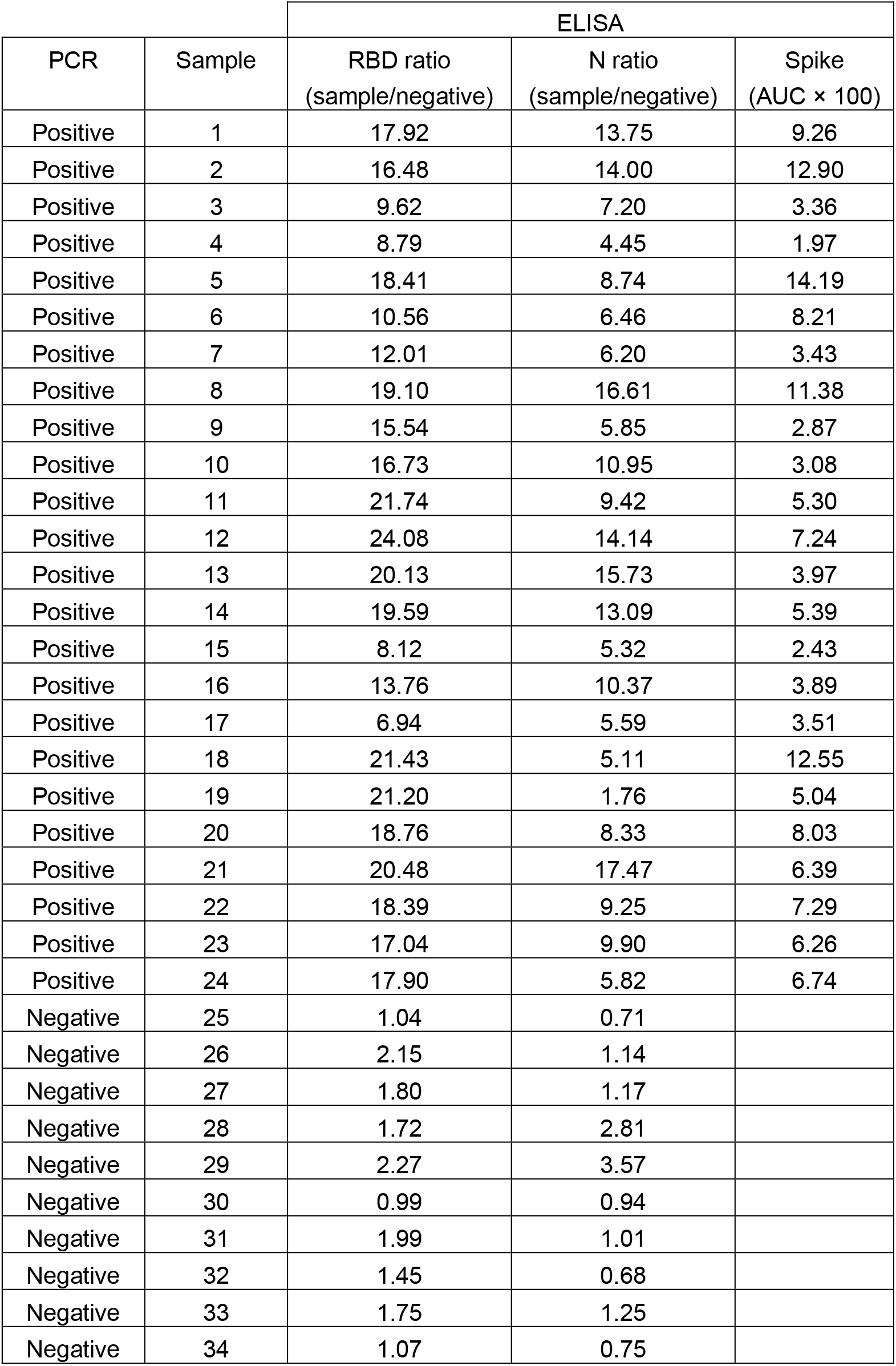
SARS-CoV-2 Protein ELISA Values

| PCR | Sample | ELISA |  |  |
| --- | --- | --- | --- | --- |
|  |  | RBD ratio<br>(sample/negative) | N ratio<br>(sample/negative) | Spike<br>(AUC × 100) |
| Positive | 1 | 17.92 | 13.75 | 9.26 |
| Positive | 2 | 16.48 | 14.00 | 12.90 |
| Positive | 3 | 9.62 | 7.20 | 3.36 |
| Positive | 4 | 8.79 | 4.45 | 1.97 |
| Positive | 5 | 18.41 | 8.74 | 14.19 |
| Positive | 6 | 10.56 | 6.46 | 8.21 |
| Positive | 7 | 12.01 | 6.20 | 3.43 |
| Positive | 8 | 19.10 | 16.61 | 11.38 |
| Positive | 9 | 15.54 | 5.85 | 2.87 |
| Positive | 10 | 16.73 | 10.95 | 3.08 |
| Positive | 11 | 21.74 | 9.42 | 5.30 |
| Positive | 12 | 24.08 | 14.14 | 7.24 |
| Positive | 13 | 20.13 | 15.73 | 3.97 |
| Positive | 14 | 19.59 | 13.09 | 5.39 |
| Positive | 15 | 8.12 | 5.32 | 2.43 |
| Positive | 16 | 13.76 | 10.37 | 3.89 |
| Positive | 17 | 6.94 | 5.59 | 3.51 |
| Positive | 18 | 21.43 | 5.11 | 12.55 |
| Positive | 19 | 21.20 | 1.76 | 5.04 |
| Positive | 20 | 18.76 | 8.33 | 8.03 |
| Positive | 21 | 20.48 | 17.47 | 6.39 |
| Positive | 22 | 18.39 | 9.25 | 7.29 |
| Positive | 23 | 17.04 | 9.90 | 6.26 |
| Positive | 24 | 17.90 | 5.82 | 6.74 |
| Negative | 25 | 1.04 | 0.71 |  |
| Negative | 26 | 2.15 | 1.14 |  |
| Negative | 27 | 1.80 | 1.17 |  |
| Negative | 28 | 1.72 | 2.81 |  |
| Negative | 29 | 2.27 | 3.57 |  |
| Negative | 30 | 0.99 | 0.94 |  |
| Negative | 31 | 1.99 | 1.01 |  |
| Negative | 32 | 1.45 | 0.68 |  |
| Negative | 33 | 1.75 | 1.25 |  |
| Negative | 34 | 1.07 | 0.75 |  |

To quantitate neutralization titers, plasma was diluted 1:100 to 1:900 and tested by authentic virus and VSV-ΔG-S GFP, Luci., and SEAP pseudotyped viruses. AUC and ND_50_ were calculated (Table 2). The average AUC values for the authentic virus neutralization assay were 51.14 (95% CI: 43.01 – 62.25) for positive participants and 10.96 (95% CI: 6.29 – 15.63) for negative participants. The GFP, Luci, and SEAP pseudotyped virus neutralization assays gave average AUC values of 71.07 (95% CI: 65.47 – 76.67), 54.20 (95% CI: 46.67 – 61.73), and 56.14 (95% CI: 48.26 – 64.01) respectively, for positive participants and 9.33 (95% CI: 3.62 – 15.04), 0 (95% CI: 0 – 0), and 1.21 (95% CI: 0 – 2.79) respectively for negative participants (Table 2). The geometric average ND_50_ value for the authentic virus neutralization assay was 254.7 (95% CI: 92.97 – 697.6) for positive participants and 13.56 (95% CI: 5.08 – 36.14) for negative participants compared to 1305 (95% CI: 763.5 – 2232), 404.1 (95% CI: 225.1 – 725.5), and 474.3 (95% CI: 255.7–879.7), for positive participants and 12.12 (95% CI: 3.562 – 41.27), 1 (95% CI: 1 – 1), and 1.772 (95% CI: 0.7476 – 4.202) for negative participants respectively for the GFP, Luci and SEAP pseudotyped viruses. All neutralization platforms differentiated average negative and positive samples (Figure 1). While the AUC and ND_50_ values were significantly higher for the GFP pseudotyped virus compared to the authentic virus or Luci. pseudotyped virus, suggesting that VSV-ΔG-S-GFP could be a more sensitive assay, it is balanced by increased AUC and ND_50_ values in negative participants. Only the Luci. and SEAP pseudotyped viruses showed no background in samples from PCR-negative participants. A Bland-Altman methods comparison test shows that there is systematic bias between the different pseudotyped virus neutralization assays and the authentic virus neutralization assay, leading to higher variability (highest for GFP pseudotypes) when the signal is low for each assay (Supplemental Figure 1). However, this bias decreases when signal becomes higher, resulting in the pseudotype assays becoming more concordant with authentic virus neutralization. The sigmoidal relationship between the amount of analyte detected and the readout in SEAP and luminescence assays could be a reason for the difference in bias between the pseudotyped virus assays. Variance at the lower end of the curve is less likely to be detected above background, compared to authentic virus neutralization and GFP pseudotyped virus neutralization where each infectious unit is counted, and variance has the same magnitude in both negative and positive samples. Furthermore, SEAP and luminescence detection kits often provide controls and stringent parameters for keeping background and noise to minimal levels.

**Table 2:** Authentic Virus and Pseudotyped Virus Neutralization Summary Statistics

| PCR | Sample | Authentic Virus |  | GFP Pseudotype |  | Luci. Pseudotype |  | SEAP Pseudotype |  |
| --- | --- | --- | --- | --- | --- | --- | --- | --- | --- |
|  |  | AUC | ND<sub>50</sub> | AUC | ND<sub>50</sub> | AUC | ND<sub>50</sub> | AUC | ND<sub>50</sub> |
| Positive | 1 | 0.565 | 380 | 0.739 | 908 | 0.520 | 276 | 0.582 | 382 |
| Positive | 2 | 0.630 | 671 | 0.804 | 2305 | 0.692 | 662 | 0.645 | 535 |
| Positive | 3 | 0.166 | 1 | 0.367 | 153 | 0.137 | 33 | 0.164 | 60 |
| Positive | 4 | 0.321 | 1 | 0.309 | 116 | 0.237 | 78 | 0.287 | 109 |
| Positive | 5 | 0.831 | 2527 | 0.860 | 5744 | 0.821 | 2162 | 0.836 | 2834 |
| Positive | 6 | 0.662 | 745 | 0.723 | 1505 | 0.668 | 692 | 0.706 | 722 |
| Positive | 7 | 0.206 | 51 | 0.661 | 672 | 0.467 | 253 | 0.412 | 181 |
| Positive | 8 | 0.794 | 2612 | 0.810 | 2997 | 0.764 | 1341 | 0.814 | 1922 |
| Positive | 9 | 0.095 | 8 | 0.613 | 665 | 0.314 | 112 | 0.322 | 121 |
| Positive | 10 | 0.284 | 86 | 0.633 | 586 | 0.412 | 193 | 0.368 | 154 |
| Positive | 11 | 0.649 | 782 | 0.822 | 3747 | 0.531 | 307 | 0.518 | 296 |
| Positive | 12 | 0.702 | 682 | 0.792 | 1995 | 0.597 | 447 | 0.645 | 527 |
| Positive | 13 | 0.646 | 606 | 0.837 | 3229 | 0.638 | 551 | 0.606 | 392 |
| Positive | 14 | 0.468 | 275 | 0.701 | 716 | 0.444 | 208 | 0.413 | 186 |
| Positive | 15 | 0.232 | 93 | 0.599 | 462 | 0.301 | 116 | 0.269 | 105 |
| Positive | 16 | 0.825 | 2769 | 0.833 | 2317 | 0.820 | 2606 | 0.848 | 2910 |
| Positive | 17 | 0.429 | 281 | 0.770 | 3222 | 0.470 | 248 | 0.493 | 259 |
| Positive | 18 | 0.886 | 35373 | 0.887 | 50378 | 0.886 | 37272 | 0.890 | 64851 |
| Positive | 19 | 0.808 | 2008 | 0.842 | 3409 | 0.694 | 662 | 0.700 | 635 |
| Positive | 20 | 0.593 | 495 | 0.733 | 983 | 0.565 | 369 | 0.634 | 589 |
| Positive | 21 | 0.414 | 175 | 0.744 | 1373 | 0.603 | 417 | 0.707 | 850 |
| Positive | 22 | 0.462 | 266 | 0.724 | 1001 | 0.610 | 444 | 0.719 | 972 |
| Positive | 23 | 0.192 | 63 | 0.636 | 390 | 0.450 | 215 | 0.423 | 197 |
| Positive | 24 | 0.414 | 190 | 0.617 | 441 | 0.370 | 163 | 0.474 | 266 |
| Negative | 25 | 0.031 | 9 | 0.151 | 40 | 0.000 | 1 | 0.000 | 1 |
| Negative | 26 | 0.035 | 3 | 0.191 | 57 | 0.000 | 1 | 0.000 | 1 |
| Negative | 27 | 0.141 | 33 | 0.035 | 10 | 0.000 | 1 | 0.000 | 1 |
| Negative | 28 | 0.212 | 54 | 0.007 | 2 | 0.000 | 1 | 0.000 | 1 |
| Negative | 29 | 0.176 | 43 | 0.051 | 15 | 0.000 | 1 | 0.060 | 18 |
| Negative | 30 | 0.000 | 1 | 0.231 | 82 | 0.000 | 1 | 0.000 | 1 |
| Negative | 31 | 0.192 | 37 | 0.000 | 1 | 0.000 | 1 | 0.060 | 17 |
| Negative | 32 | 0.110 | 18 | 0.209 | 68 | 0.000 | 1 | 0.000 | 1 |
| Negative | 33 | 0.051 | 4 | 0.000 | 1 | 0.000 | 1 | 0.000 | 1 |
| Negative | 34 | 0.148 | 38 | 0.059 | 18 | 0.000 | 1 | 0.000 | 1 |

**Figure 1:**
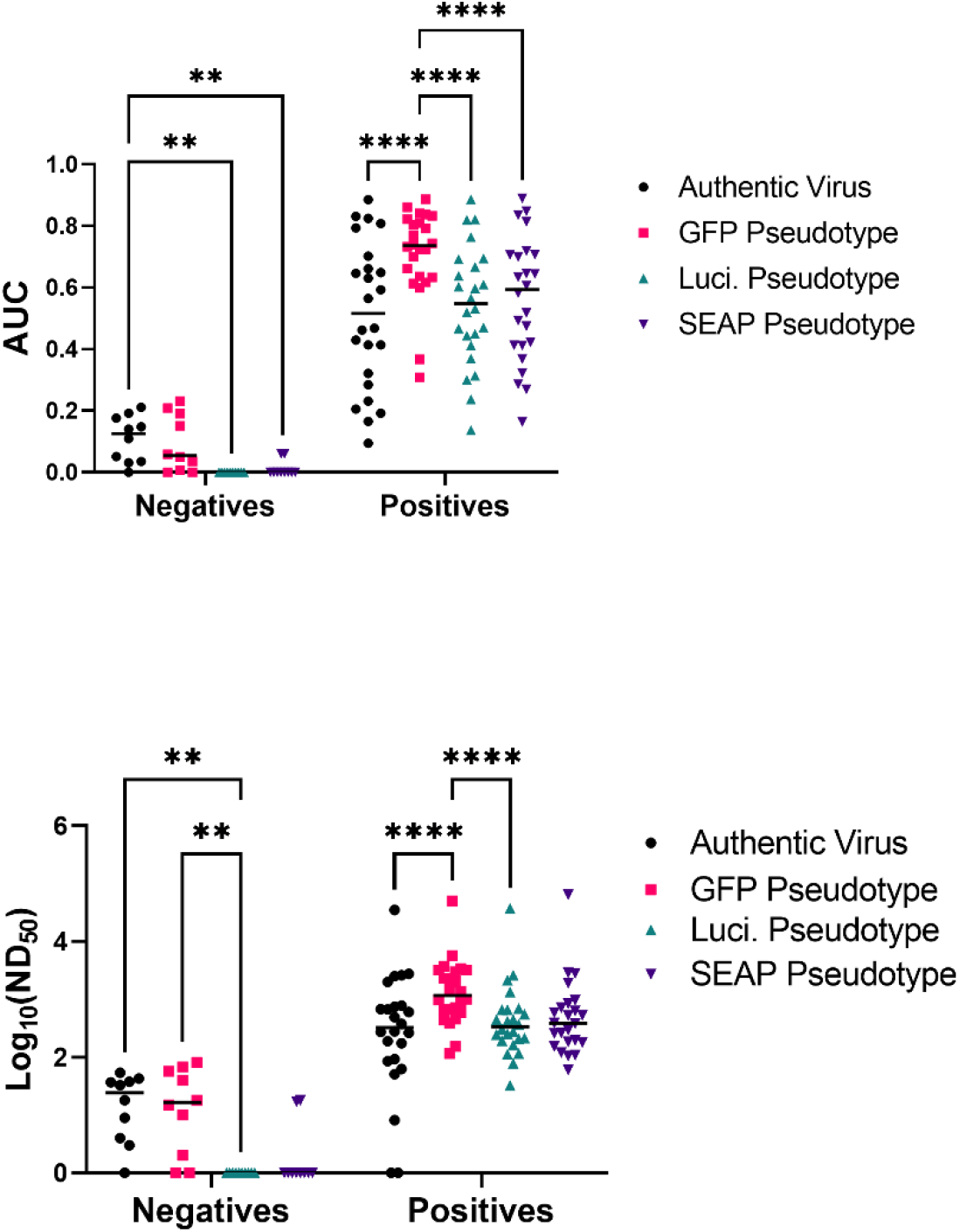
Comparison of neutralization assays by sample groups. Area under the curve (AUC) (top) and neutralization dilution – 50% (ND_50_) (bottom) calculations by neutralization assay type. AUC and ND_50_ values were calculated and used to compare authentic virus neutralization (black), GFP pseudotype neutralization (pink), luciferase pseudotype (teal), and SEAP pseudotype (purple). * p < 0.05, ** p < 0.01, ***p < 0.001 (mixed-effects model with the Geisser-Greenhouse correction and Tukey multiple comparisons post-test and p-value adjustment). n = 34 samples run on each assay.

To determine which neutralization platforms best correlated with authentic virus, linear regression analyses were performed. All pseudotyped virus neutralization platforms were significantly correlated to authentic virus neutralization regardless of the reporter with Luci. (Pearson’s r = 0.765) and SEAP (Pearson’s r = 0.775) having the highest correlations (Figure 2A). The pseudotyped virus neutralization assays were significantly correlated with each other with Pearson’s r values as high as 0.971 between the Luci. and SEAP assays. Linear regression analyses demonstrated that ELISA titers to the RBD (Pearson’s r = 0.691) and spike (Pearson’s r = 0.648) are also significantly correlated with authentic virus neutralization titers (Figure 2B). Nucleocapsid ELISA was significantly correlated with authentic virus neutralization, but has the worst correlation with authentic virus neutralization (Pearson’s r = 0.514), which has been shown previously for pseudotyped virus neutralization^16^. This is also congruent with the observation that antibodies targeting the RBD domain of spike are highly neutralizing^17^. A principle component analysis (PCA) was performed using all three ELISAs and all three pseudotyped virus platforms as variables (Supplemental Figure 2). The resulting PCA plots shows distinct clustering of the samples with the highest authentic virus neutralization titers and a gradient from poorly neutralizing samples (in the bottom left) to highly neutralizing samples (in the top right). Finally, the average difference between the log(ND_50_) for authentic virus neutralization and each pseudotyped virus neutralization is −0.487 for the GFP pseudotype, 0.191 for the Luci. pseudotype, and 0.069 for the SEAP pseudotype.

**Figure 2:**
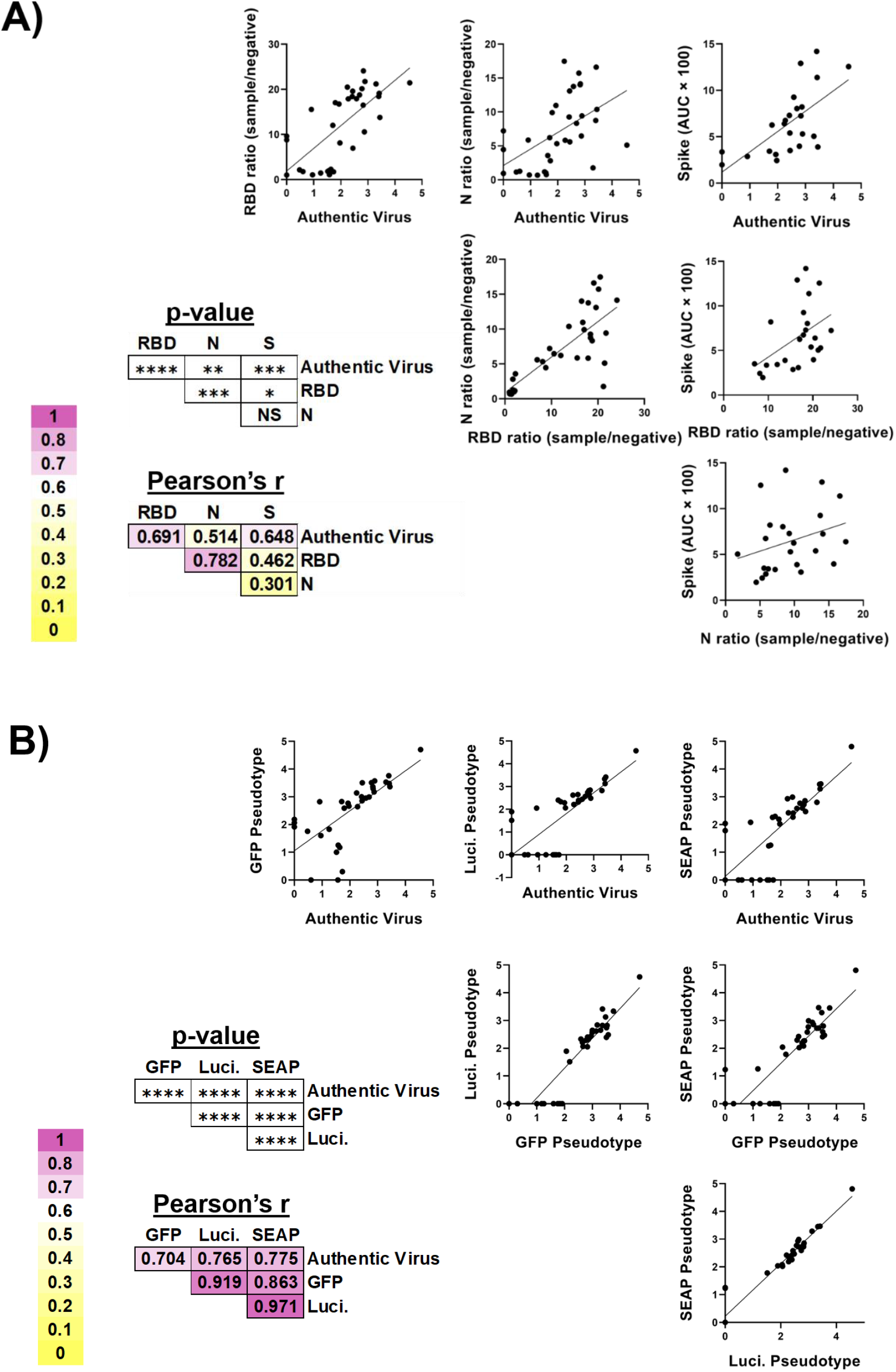
Correlation of SARS-CoV-2 serologic assays. A) SARS-CoV-2 specific ELISA assays and B) VSV pseudotyped virus neutralization assays were compared by simple linear regression. The Pearson’s r values (a metric of correlation) and p-values corresponding to each graph are to the lower left of each set of graphs. The shade of background corresponds to the degree of correlation between the two assays. * p < 0.05, ** p < 0.01, *** p < 0.001, **** p < 0.0001.

To assess granularity in the different ELISA results, cut-off values were used to categorize responses as high positive, low positive, or negative. Determination of cut-off values (RBD ratio: 15, nucleocapsid ratio:10, and spike: 6) was done by finding internal nadir present in histograms for the different ELISAs. The stratification of RBD ELISA responses into high and low groups did not result in significantly different responses in any of the neutralization assays (Figure 3A), suggesting that high RBD values do not necessarily correlate to higher neutralization titers, despite RBD ELISA positivity being associated with neutralization (Figure 2A). Similar results were obtained for the spike ELISA (Figure 3B) and nucleocapsid ELISA (Figure 3C). There was, however, a trend for increased neutralization in the high positive group versus the low positive group for each neutralization assay, regardless of ELISA assay, justifying future studies specifically designed to test the granularity of these assays. Congruent with the findings in Figure 2, highly positive ELISA results were significantly better at neutralizing than the negative samples for each ELISA.

**Figure 3:**
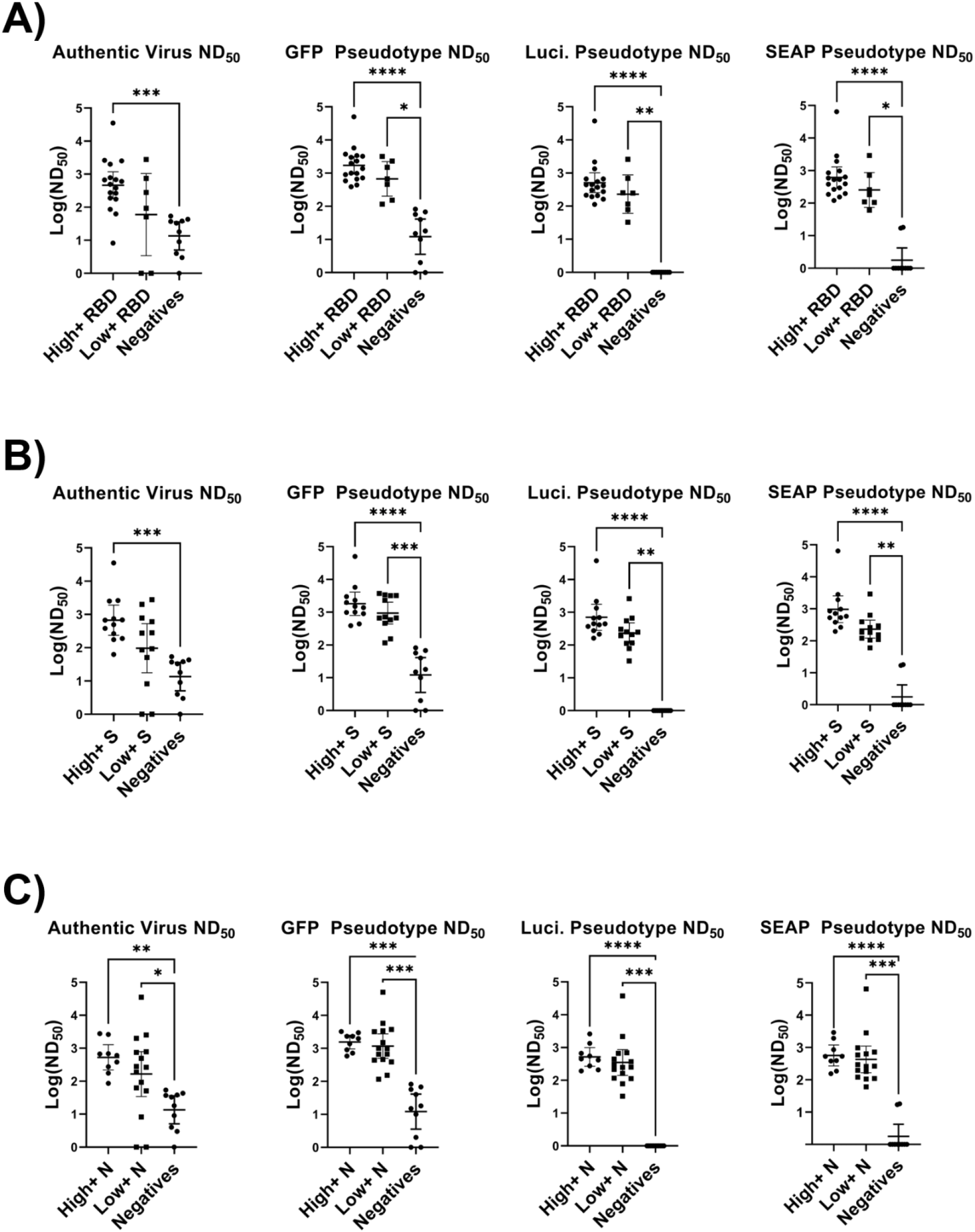
Comparison of high positive, low positive, and negative ELISA groups across neutralization assays. A) RBD B) spike (S) and C) nucleocapsid (N) positive samples were divided into high and low positives by finding cut off values using histograms (RBD ratio: 15, N ratio: 10, and Spike: 6). Log(ND_50_) values for the corresponding samples were then graphed and compared by Kruskal-Wallis test with Dunn’s multiple comparisons tests. Significance thresholds: * p < 0.05, ** p < 0.01, *** p < 0.001, p **** p < 0.0001.

While all the serological assays were significantly correlated with authentic virus neutralization, some assays performed better than others at predicting authentic virus neutralization (Supplemental Table 2). Based on correlation with authentic virus neutralization, the most accurate assays were the Luci. and SEAP pseudotyped virus neutralization assays. GFP pseudotyped virus neutralization, spike ELISA, and RBD ELISA form a second tier of assays which are still quite accurate at predicting authentic virus neutralization. Furthermore, the GFP pseudotyped virus neutralization was able to detect antibodies at significantly higher dilutions than the other assays, making it the most sensitive assay tested. Despite nucleocapsid antigen being the basis for several common commercial antibody tests, nucleocapsid was the least predictive of authentic virus neutralization.

Collectively, these data demonstrate that VSV-ΔG pseudotyped virus neutralization platforms, especially Luci. and SEAP based platforms, are better at predicting authentic virus neutralization than ELISA regardless of the viral antigen tested. Not only are the Luci. and SEAP based pseudotype platforms most strongly correlated to authentic virus neutralization, they also have the lowest average difference in log(ND_50_) compared to authentic virus neutralization. Previous reports have only compared ELISA titers to pseudotyped virus neutralization^16^, ELISA to authentic virus neutralization^18^, or only one type of ELISA and one pseudotyped virus platform against authentic virus neutralization^10^. Our studies provide one of the most comprehensive comparisons amongst multiple ELISA antigens, pseudotyped virus neutralization platforms, and authentic virus neutralization.

Of note, several spike and RBD positive samples showed very little authentic virus neutralization despite having moderate to high neutralization on the pseudotyped virus platforms. Furthermore, one sample appeared to show antibody dependent enhancement (ADE) in the authentic virus neutralization assay (1.8-fold increased plaque forming units (PFU)), but still showed low but detectable neutralization in all the pseudotyped virus platforms. While there is no definitive role for ADE during human SARS-CoV-2 infection, ADE has been demonstrated *in vitro* with other human coronaviruses^19^. Further characterization of this sample and screening for and characterizing similar samples will lead to a better understanding of the risk of ADE during SARS-CoV-2 infection. Recent evidence suggests that several SARS-CoV-2 variants, including B.1.351 and P.1, have decreased neutralization when treated with monoclonal antibodies or polyclonal sera derived from patients infected with early strains of SARS-CoV-2^20-22^. Future studies need to assess how the mutations present in the variants differentially affect ELISA, pseudotyped virus neuralization, and authentic virus neutralization.

In addition to accuracy, the serological assays differ in several key features (Supplemental Table 2), and the assay of choice may have to be determined by the settings. The requirement for a BSL3 laboratory makes authentic virus assays technically challenging and unfeasible for many clinical and research applications. This can be overcome by pseudotyped viruses. However, creation and validation of the different pseudotyped viruses is not trivial and read-outs may require specialized equipment (e.g. luminometer for the Luci. platform). Most laboratories have ready access to the equipment needed for performing ELISAs, making the technical requirements for these assays low. ELISAs can also be completed within several hours while the pseudotyped virus neutralization platforms require 12-24-hour incubations and authentic virus neutralization requires 48-72 hours. If all technical requirements have been met and are available, the assays are all relatively inexpensive, except for the Luci. platform which requires expensive reagents for reading the results. If turnaround time is a priority, the RBD and spike ELISAs would provide the fastest results with minor decreases in predicting authentic virus neutralization response. Alternatively, in resource-limited settings like field hospitals, the GFP based pseudotyped virus neutralization assay requires only a basic fluorescence microscope for readout and is more predictive of authentic virus neutralization than any of the ELISAs. Overall, this study shows that all six serological assays, to varying degrees, correlated with authentic virus neutralization, and the optimal serological assay for assessing a protective antibody response is going to be institution and question specific.

## Data Availability

All data is available in the manuscript.

## Methods

### Data Availability

Authors can confirm that all relevant data are included in the paper and/or its supplementary information files.

### RBD/N ELISA

SARS-CoV-2 RBD protein was diluted to a concentration of 1.5 µg/ml in PBS and added at 50 µl per well to a 96-well ELISA plate. The ELISA plates were sealed and allowed to incubate at 4°c overnight. The next day the coating solution was removed, and the plates were blocked at room temperature (RT) using 3% milk (200 µl per well) for a minimum of 1 hour but not exceeding 4 hours. While the plates were being blocked, the samples were prepared by diluting the plasma 1:50 in 1% milk. Following the blocking period, the milk was removed, and the plates were washed 3x with 0.1% phosphate buffered saline containing 0.1% Tween-20 (PBS-T) using 200 µl per well. The diluted plasma was added to the blocked plate at 50 µl per well along with 2 positive controls (α SARS-CoV-2 RBD antibody at 1:5000, 1:25000, 1:125000, and 1:625000 dilutions and plasma from a naturally infected donor at a 1:50 dilution) and a known negative, pre-pandemic plasma sample (1:50). The samples were incubated for 1.5 hours at RT and then removed and washed 3x with 200 µl 0.1% PBS-T. Goat α human IgG horseradish peroxidase (HRP) conjugated secondary antibody was diluted 1:2500 in 1% milk and 50 µl was added to each well of the washed plate and incubated at RT for 30 minutes. Following the incubation period, the secondary was removed, and the plate was washed 3x with 0.1% PBS-T. OPD substrate was prepared directly before use and added at 50 µl per well for exactly 8 minutes. The O-phenylenediamine dihydrochloride (OPD) substrate was stopped by adding 50 µl of 3M HCl and then the plate was read using a spectrophotometer at 490nm.

### Spike ELISA

SARS-CoV-2 spike protein was diluted to a concentration of 2 µg/ml in PBS and added at 50 µl per well to a 96-well ELISA plate. The ELISA plates were sealed and allowed to incubate at 4°c overnight. The next day the coating solution was removed, and the plates were blocked using 3% milk (200 µl per well) for a minimum of 1 hour but not exceeding 4 hours. While the plates were being blocked, the samples were prepared by creating a 3-fold serial dilution starting at 1:100 and ending at 1:8100 (1% milk as diluent). Following the blocking period, the milk was removed, and the plates were washed 3x with 0.1% PBS-T using 200 µl per well. The diluted plasma was added to the blocked plate at 50 µl per well along with 2 positive controls (α SARS-CoV-2 RBD antibody at 1:5000, 1:25000, 1:125000, and 1:625000 dilutions and plasma from a naturally infected donor at 1:100, 1:300, 1:900, 1:2700, and 1:8100 dilutions) and a known negative, pre-pandemic plasma sample (1:100). The samples were incubated for 1.5 hours at RT and then removed and washed 3x with 200 µl 0.1% PBS-T. Goat α human IgG HRP conjugated secondary antibody was diluted 1:2500 in 1% milk and 50 µl was added to each well of the washed plate and incubated at RT for 30 minutes. Following the incubation period, the secondary was removed, and the plate was washed 3x with 0.1% PBS-T. OPD substrate was prepared directly before use and added at 50 µl per well for exactly 8 minutes. The OPD substrate was stopped by adding 50 µl of HCL acid and then the plate was read using a spectrophotometer at 490nm. Spike data is presented as either AUC or AUC × 100 in order to plot it on the same scale as the other ELISAs.

### Tissue culture

VeroE6 cells stably expressing TMPRSS2 (Vero-TMPRSS2) (XenoTech) were cultured in Eagle’s Minimal Essential Medium (EMEM) supplemented with 10% fetal bovine serum (FBS), 100 U/mL penicillin, 100 µg/mL streptomycin, and 2 mM GlutaMax (Gibco). Media was supplemented with 1 mg/mL G418 every other passage. All tissue culture was performed in a humidified incubator set to 37° C and 5% CO_2_.

### SARS-CoV-2 neutralizing antibody assay

Serially diluted plasma samples were mixed with diluted (approximately 6 PFU/cm2) SARS-CoV-2 (2019n-CoV/USA_WA1/2020) in EMEM supplemented with 5% FBS, 100 U/mL penicillin, 100 µg/mL streptomycin, and 2 mM GlutaMax. Mixtures were incubated for 1 h in a humidified incubator at 37° C and 5% CO_2_. After 1 h, culture media was removed from approximately 90% confluent Vero-TMPRSS2 cells grown in 6-well plates and replaced with virus/plasma mixtures. Plates were returned to the incubator for 1 h at 37° C and 5% CO_2_.

Plates were rocked manually every 15 minutes. After incubation, an agarose overlay containing Minimal Essential Media (MEM) supplemented with 5% FBS 100 U/mL penicillin, 100 µg/mL streptomycin, 2 mM GlutaMax, 0.075% sodium bicarbonate, 0.01 M 4-(2-hydroxyethyl)-1-piperazineethanesulfonic acid (HEPES), and 1% low melting temperature agarose (SeaPlaque; Lonza) was added to each well. Once agarose hardened at RT, plates were returned to the incubator at 37° C and 5% CO_2_. After 48 h, cells were fixed with 10% neutral buffered formalin for 1 h, the agar plugs were removed, and then cells were stained with crystal violet for 5 – 10 minutes. Upon rinsing with H20, plaques were visualized and counted. All samples were run in duplicate

### VSV-ΔG-GFP-SARS-CoV-2-S Neutralizing antibody assay

Serially diluted plasma samples were mixed with diluted and mixed with Spike/VSV-ΔG-GFP pseudotypes in EMEM supplemented with 5% FBS, 100 U/mL penicillin, 100 µg/mL streptomycin, and 2 mM GlutaMax. Mixtures were incubated for 1 h in a humidified incubator at 37° C and 5% CO_2_. After 1 h, culture media was removed from approximately 90% confluent Vero-TMPRSS2 cells grown in 96-well plates and replaced with virus/plasma mixtures. Plates were returned to the incubator at 37° C and 5% CO_2_. After 24 h, IU were quantified manually using an EVOS fluorescence microscope. All samples run in duplicate.

### Luciferase Assay

20 hours prior to assay set up, Vero-TMRSS2 were plated in a 96-well plate at 20,000 cells per well in DMEM supplemented with 5% FBS and 1 mg/mL G418. For assay set up, plasma samples were initially diluted 1:100 and serially diluted 1:3 in DMEM supplemented with 5% FBS. Diluted samples were mixed 1:1 with Spike/VSV-ΔG-Luciferase pseudotyped virus diluted to final 250 IU per well in serum free DMEM. Mixtures were incubated for 1 hour in a humidified incubator at 37° C and 5% CO_2_. After the incubation period, culture medium was removed from Vero-TMPRSS2 cells and virus/plasma mixture was added to the cells in triplicate. Plates were incubated for approximately 18 hours in a humidified incubator at 37° C and 5% CO_2_. After the incubation period, Luc-Screen Extended-Glow (ThermoFisher) buffers were added to the wells according to manufacturer’s instructions and incubated for a minimum of 10 minutes at room temperature protected from light. Luminescence was measured with a luminometer using one second integration time.

### SEAP Assay

20 hours prior to assay set up, Vero-TMRSS2 were plated in a 96-well plate at 20,000 cells per well in DMEM supplemented with 5% FBS and 1 mg/mL G418. For assay set up, plasma samples were initially diluted 1:100 and serially diluted 1:3 in DMEM supplemented with 5% FBS. Diluted samples were mixed 1:1 with purified Spike/VSV-ΔG╌SEAP pseudotyped virus diluted to final 250 IU per well in serum free DMEM. Mixtures were incubated for 1 hour in a humidified incubator at 37° C and 5% CO_2_. After the incubation period, culture medium was removed from Vero-TMPRSS2 cells and virus/plasma mixture was added to the cells in triplicate. Plates were incubated for approximately 28 hours in a humidified incubator at 37° C and 5% CO_2_. After the incubation period, Quanti-Blue (InvivoGen) solution was combined with 20 μL supernatant according to manufacturer’s instructions and incubated for a minimum of 15 minutes at 37° C protected from light. Optical density was measured at 620-655 nm.

### SARS-CoV-2/VSV pseudotype production

VSV-ΔG pseudotypes displaying the full-length SARS-CoV-2 spike (Wuhan-Hu-1 strain) were generated essentially as described^23^ with the following modifications. Baby hamster kidney (BHK-21) cells in 10 cm dishes were transfected using Lipofectamine 2000 according to the manufacturer’s instructions with 24 μg of a plasmid encoding a codon-optimized cDNA for the SARS-CoV-2 spike^15^, which was generously provided by Florian Krammer. Approximately 20-24 hours later the transfected cells were infected at a multiplicity of 5 with VSV-G pseudotyped ΔG-GFP, luciferase, or SEAP. Virus was adsorbed for 1 hr, the inoculum was removed, cells were rinsed once with serum-free DMEM and then 4 ml of hybridoma supernatant containing the I1 monoclonal antibody^24^ was added for 30 minutes to neutralize residual VSV-ΔG pseudotyped virus from the inoculum and then replaced with DMEM containing 20% fetal bovine serum. The supernatant containing the spike-ΔG pseudotypes was collected 22-24 hours later, cell debris was removed by centrifugation at 450 x g for 10 minutes. For the ΔG-GFP and luciferase pseudotypes, the supernatant was aliquoted and stored at −80° C. For the ΔG-SEAP pseudotypes, the supernatant was transferred to a Beckman SW41 tube, underlayered with sterile 20% sucrose in PBS, and virus was pelleted at 35,000 rpm for 45 minutes is a SW41 swinging bucket rotor. Pelleting virus was required to separate it from SEAP released from the infected cells. The pellets were resuspended in DMEM containing 20% FBS and stored at −80° C.

### Statistics

Area under the curve (AUC) and neutralization dilution – 50% (ND_50_) analyses were performed in GraphPad Prism (version 9.0.0): non-linear regression (dose-response). Pearson’s r values for comparing assays by percent maximum AUC were calculating using simple linear regression analysis in GraphPad Prism. AUC and ND_50_ values for the different assays were compared by mixed-effects model with the Geisser-Greenhouse correction and Tukey multiple comparisons post-test and p-value adjustment in GraphPad Prism (version 9.0.0). Kruskal-Wallis tests with Dunn’s multiple comparisons tests. were performed to compare neutralizing antibody responses between highly positive ELISA samples, low positive ELISA samples, and negative samples. Principle component analysis (PCA) was performed in GraphPad Prism (version 9.0.0) with principle components selected based on parallel analysis. A 95% percentile level was used, and 1000 simulations were performed for the PCA. The Bland-Altman analyses were performed in GraphPad Prism (version 9.0.0).

## Acknowledgements

The authors wish to thank Amy E. Davis, Virginia Hargest, Rebekah Honce, Brandi Livingston, Victoria Meliopoulos, Bridgett Sharp, Maria Smith, and Kristin Wiggins for aiding the Schultz-Cherry lab’s COVID-19 response; the members of the Thomas, McGargill, and Schultz-Cherry labs for technical assistance and feedback on the work; Dr. Gang Wu and the Center for Applied Bioinformatics at St. Jude along with Michael Meagher, Timothy Lockey, and the St. Jude Good Manufacturing Practice (GMP) facility; Tamanna Shamrin and Rishi Kodela for creation and management of the clinical database. This work was funded by ALSAC, the NIAID for the St. Jude Center of Excellence for Influenza Research and Surveillance (CEIRS contract HHSN27220140006C) and the NIAID Collaborative Influenza Vaccine Innovation Centers (CIVIC) contract 75N93019C00052, and by the University of Tennessee Research Foundation. JHE is supported by the American Society of Hematology Scholar Award. Work in the Krammer laboratory is partially funded by the NIAID Collaborative Influenza Vaccine Innovation Centers (CIVIC) contract 75N93019C00051, NIAID Center of Excellence for Influenza Research and Surveillance (CEIRS, contract # HHSN272201400008C), by the generous support of the Cohen Foundation, the JPB Foundation and the Open Philanthropy Project (research grant 2020-215611 (5384), and by anonymous donors.

## Author contributions

Conceptualization: N.W., K.W., S.C., E.K.R., S.S.-C.

Conception, design, and oversight of parent study; SJTRC: J.H., H.H., K.J.A., R.D., A.H.G., J.H.E., J.W., and P.G.T.

Formal Analysis: NW., K.W., S.C., E.K.R., T.M., L.T.

Investigation: N.W., K.W., S.C., E.K.R.

Methodology: NW., K.W., S.C., E.K.R., C.Y.L. E.K.A., P.F., M.A.M.

Resources: F.K., M.A.W.

Sample acquisition and curation: K.J.A., A.G., A.H.G., E.K.A., J.H.E., J.W., and P.G.T. Writing, original draft: N.W., E.K.R.

Writing, review, and editing: N.W., K.W., S.C., E.K.R., M.A.W., H.H., J.H.E, A.H.G., T.M., T.L., D.R.H., M.A.M., F.K., J.W., P.G.T., S.S.-C.

Visualization: N.W.

Supervision: S.S.-C.

