## Supplementary material for "Choosing the right tool for the job: A comprehensive assessment of serological assays for SARS-CoV-2 as surrogates for authentic virus neutralization": Supplemenal figures

**Supplemental Data**

Nicholas Wohlgemuth[1], Kendall Whitt[2], Sean Cherry[1], Ericka Kirkpatrick Roubidoux[1], Chun-Yang Lin[3], Kim J. Allison[1], Ashleigh Gowen[1], Pamela Freiden[1], E. Kaitlynn Allen[3], St. Jude Investigative Team, Aditya H. Gaur[1], Jeremie H. Estepp[4,5], Li Tang[6], Tomi Mori[6], Diego R. Hijano[1], Hana Hakim[1], Maureen A. McGargill[3], Florian Krammer[7], Michael A. Whitt[2], Joshua Wolf[1], Paul G. Thomas[3], Stacey Schultz-Cherry[1*]

St. Jude Investigative Team

David C. Brice[3], Ashley Castellaw[3], David E. Whittman[8], Jason Hodges[4], Ronald H. Dallas[1], Valerie Cortez[1], Ana Vazquez-Pagan[1], Richard J. Webby[1], Resha Bajracharya[3], Brandi L Clark[3], Lee-Ann Van de Velde[3], Walid Awad[3], Taylor L Wilson[3], Allison M. Kirk[3], Elaine I. Tuomanen[1], Richard J. Webby[1], Randall T. Hayden[9], James Hoffman[10], Jamie Russell-Bell[1], James Sparks[5]

[1]Department of Infectious Diseases, St Jude Children's Research Hospital, Memphis, TN, USA

[2]Department of Microbiology, Immunology and Biochemistry, University of Tennessee Health Science Center, Memphis, TN, USA

[3]Department of Immunology, St Jude Children's Research Hospital, Memphis, TN, USA

[4]Department of Hematology, St Jude Children's Research Hospital, Memphis, TN, USA

[5]Department of Global Pediatric Medicine, St Jude Children's Research Hospital, Memphis, TN, USA

[6]Department of Biostatistics, St Jude Children's Research Hospital, Memphis, TN, USA

[7]Department of Microbiology, Icahn School of Medicine at Mount Sinai, New York, NY, USA

[8]Office of Quality and Patient Care St Jude Children's Research Hospital, Memphis, TN, USA

[9]Department of Pathology, St Jude Children's Research Hospital, Memphis, TN, USA

[10]Department of Pharmaceutical Sciences, St Jude Children's Research Hospital, Memphis, TN, USA

Supplemental Table 1: Participant characteristics

| | No SARS-CoV-2 infection (n = 10) | | SARS-CoV-2 Infection (n = 24) | |
|---|---|---|---|---|
| **Participant characteristics*** | **n** | **(%)** | **n** | **(%)** |
| **Age in years, median (IQR)** | 48.5 | (32.5;57) | 45.5 | (38;57) |
| **Gender** | | | | |
| Female | 6 | (60%) | 20 | (83.3%) |
| Male | 4 | (40%) | 4 | (16.7%) |
| **Race** | | | | |
| White, Caucasian | 9 | (90%) | 16 | (66.7%) |
| Black, African American | 1 | (10%) | 7 | (29.2%) |
| Other | 0 | (0%) | 1 | (4.2%) |
| **Ethnicity** | | | | |
| Hispanic | 0 | 0% | 2 | (8.3%) |
| Non-Hispanic | 10 | (100%) | 22 | (91.7%) |

***All participant characteristics self-reported**

Supplemental Table 2: Logistical Attributes of SARS-CoV-2 Serological Assays

| Assay | Accuracy | Technical Requirements | Assay Time | Price | Detection Method |
|---|---|---|---|---|---|
| RBD ELISA | ++ | + | + | $ | Enzymatic reaction |
| N ELISA | + | + | + | $ | Enzymatic reaction |
| Spike ELISA | ++ | + | + | $ | Enzymatic reaction |
| Authentic Virus Neutralization | +++ | +++ | +++ | $ | Infectious unit |
| GFP Pseudotype Neutralization | ++ | ++ | ++ | $ | Infectious unit |
| Luci. Pseudotype Neutralization | +++ | ++ | ++ | $$$ | Enzymatic reaction |
| SEAP Pseudotype Neutralization | +++ | ++ | ++ | $ | Enzymatic reaction |

The number of symbols (+ or $) is a relative estimate of the column variable. Price indicates the price of running the assay given access to all the technical requirements for the assay.

Supplemental Figure 1

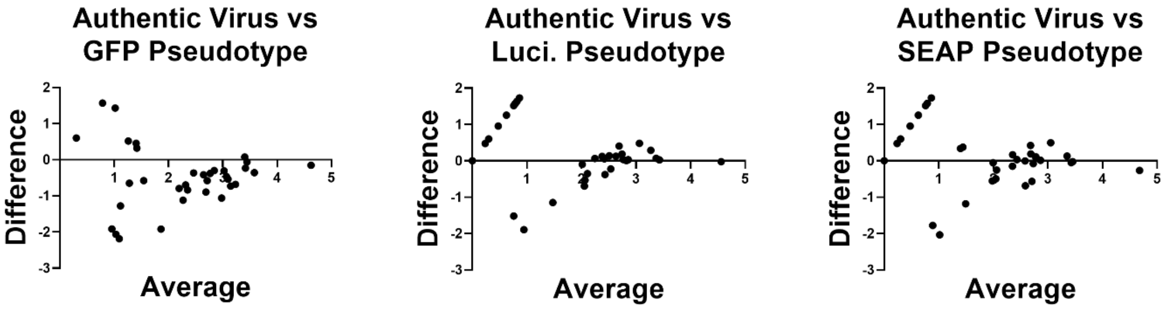


**Supplemental Figure 1: Bland-Altman analysis of SARS-CoV-2 pseudotyped virus neutralization assays**. Bland-Altman analysis was performed between the log($ND_{50}$) values of authentic virus neutralization against each pseudotyped virus platform. The difference between the two assays for each sample is on the y-axis, and the average of the two assays is on the x-axis.

Supplemental Figure 2

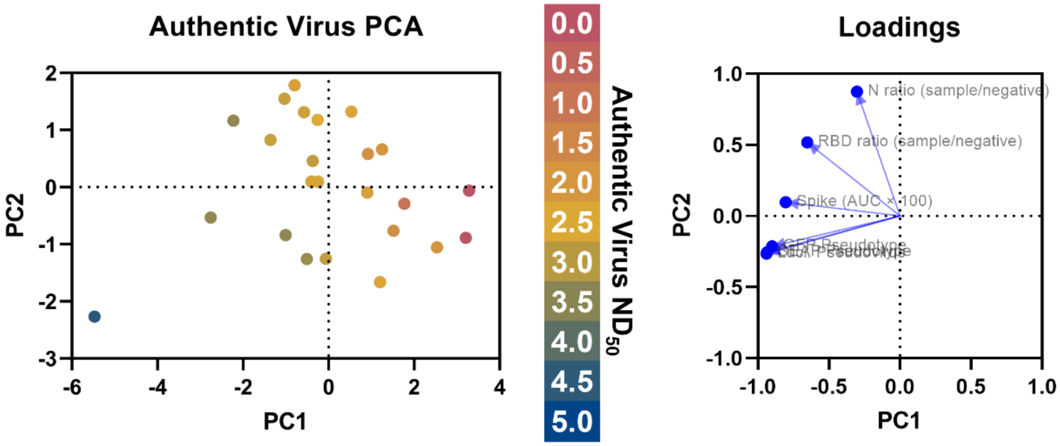


**Supplemental Figure 2: Principle component analysis (PCA) of SARS-CoV-2 serological assays**. Principle component analysis was performed using all three ELISAs (spike, RBD, and nucleocapsid) and pseudotyped virus neutralization platforms (GFP, Luci. and SEAP). The authentic virus $ND_{50}$ is indicated by the color of the data point. PCA loadings generated during the analysis are shown on the right.
